# Combining Asian-European Genome-Wide Association Studies of Colorectal Cancer Improves Risk Prediction Across Race and Ethnicity

**DOI:** 10.1101/2023.01.19.23284737

**Authors:** Minta Thomas, Yu-Ru Su, Elisabeth A. Rosenthal, Lori C Sakoda, Stephanie L Schmit, Maria N Timofeeva, Zhishan Chen, Ceres Fernandez-Rozadilla, Philip J Law, Neil Murphy, Robert Carreras-Torres, Virginia Diez-Obrero, Franzel JB van Duijnhoven, Shangqing Jiang, Aesun Shin, Alicja Wolk, Amanda I Phipps, Andrea Burnett-Hartman, Andrea Gsur, Andrew T Chan, Ann G Zauber, Anna H Wu, Annika Lindblom, Caroline Y Um, Catherine M Tangen, Chris Gignoux, Christina Newton, Christopher A. Haiman, Conghui Qu, D Timothy Bishop, Daniel D Buchanan, David R. Crosslin, David V Conti, Dong-Hyun Kim, Elizabeth Hauser, Emily White, Erin Siegel, Fredrick R Schumacher, Gad Rennert, Graham G Giles, Heather Hampel, Hermann Brenner, Isao Oze, Jae Hwan Oh, Jeffrey K Lee, Jennifer L Schneider, Jenny Chang-Claude, Jeongseon Kim, Jeroen R Huyghe, Jiayin Zheng, Jochen Hampe, Joel Greenson, John L Hopper, Julie R Palmer, Kala Visvanathan, Keitaro Matsuo, Koichi Matsuda, Keum Ji Jung, Li Li, Loic Le Marchand, Ludmila Vodickova, Luis Bujanda, Marc J Gunter, Marco Matejcic, Mark A Jenkins, Martha L Slattery, Mauro D’Amato, Meilin Wang, Michael Hoffmeister, Michael O Woods, Michelle Kim, Mingyang Song, Motoki Iwasaki, Mulong Du, Natalia Udaltsova, Norie Sawada, Pavel Vodicka, Peter T Campbell, Polly A Newcomb, Qiuyin Cai, Rachel Pearlman, Rish K Pai, Robert E Schoen, Robert S Steinfelder, Robert W Haile, Rosita Vandenputtelaar, Ross L Prentice, Sébastien Küry, Sergi Castellví-Bel, Shoichiro Tsugane, Sonja I Berndt, Soo Chin Lee, Stefanie Brezina, Stephanie J Weinstein, Stephen J Chanock, Sun Ha Jee, Sun-Seog Kweon, Susan Vadaparampil, Tabitha A Harrison, Taiki Yamaji, Temitope O Keku, Veronika Vymetalkova, Volker Arndt, Wei-Hua Jia, Xiao-Ou Shu, Yi Lin, Yoon-Ok Ahn, Zsofia K Stadler, Bethany Van Guelpen, Cornelia M Ulrich, Elizabeth A Platz, John D Potter, Christopher I Li, Reinier Meester, Victor Moreno, Jane C Figueiredo, Graham Casey, Iris Landorp Vogelaar, Malcolm G Dunlop, Stephen B Gruber, Richard B Hayes, Paul D P Pharoah, Richard S Houlston, Gail P Jarvik, Ian P Tomlinson, Wei Zheng, Douglas A Corley, Ulrike Peters, Li Hsu

**Author notes:** **Corresponding Authors:** Li Hsu: 1100 Fairview Ave. N., M2-B500, Fred Hutchinson Cancer Center, Seattle, WA 98109.; (P). Ulrike Peters: 1100 Fairview Ave. N., M4-B402, Fred Hutchinson Cancer Center, Seattle, WA 98109.; (P) 206-667- 2450.

## Abstract

Polygenic risk scores (PRS) have great potential to guide precision colorectal cancer (CRC) prevention by identifying those at higher risk to undertake targeted screening. However, current PRS using European ancestry data have sub-optimal performance in non-European ancestry populations, limiting their utility among these populations. Towards addressing this deficiency, we expanded PRS development for CRC by incorporating Asian ancestry data (21,731 cases; 47,444 controls) into European ancestry training datasets (78,473 cases; 107,143 controls). The AUC estimates (95% CI) of PRS were 0.63(0.62-0.64), 0.59(0.57-0.61), 0.62(0.60-0.63), and 0.65(0.63-0.66) in independent datasets including 1,681-3,651 cases and 8,696-115,105 controls of Asian, Black/African American, Latinx/Hispanic, and non-Hispanic White, respectively. They were significantly better than the European-centric PRS in all four major US racial and ethnic groups (p-values<0.05). Further inclusion of non-European ancestry populations, especially Black/African American and Latinx/Hispanic, is needed to improve the risk prediction and enhance equity in applying PRS in clinical practice.

## Introduction

Colorectal cancer (CRC) is a leading cause of cancer death, yet it is among the most preventable cancers via screening^1^. Together with detection of CRC at early stages, which dramatically improves prognosis, optimal screening has the potential for major impact on CRC mortality. However, current screening programs are primarily age and family-history based and more refinement through risk-based screening recommendations could be instrumental in improving their effectiveness.

Genetics plays a key role in the CRC development and, as for most cancers and other common diseases, the risk is polygenic^2^. As such, we can utilize the polygenic risk structure to develop a polygenic risk score (PRS) to quantify an individual’s inherited risk of developing CRC. As the predictive performance improves, a PRS can become clinically useful as a risk stratification tool for targeted screening and chemoprevention. However, PRS built based on European ancestry data have sub-optimal performance in other ancestral populations^3^ because of differential linkage disequilibrium (LD) patterns and allele frequencies across racial and ethnic groups for disease risk variants of CRC^4–9^. The poor transferability of PRS across racial and ethnic groups has raised concern regarding whether its application in clinical practice may exacerbate existing health disparities^7^. As a result, there is a need to improve the accuracy of polygenic prediction across different racial and ethnic groups to maximize the clinical and public-health translational potential of PRS and enhance equity in precision medicine.

Developing ancestry-specific PRS requires sufficient sample sizes for each ancestral group; however, the sample sizes for non-European ancestry groups, while increasing, remain only a fraction of the sample size for European ancestry. Existing studies suggest that leveraging information from other ancestries can improve ancestry-specific PRS ^10,11^. As an alternative to developing ancestry-specific PRS, one may develop a single cross-ancestry PRS based on meta-analysis of genome-wide association studies (GWAS) across all available ancestral groups^12–14^. To our knowledge, there is no study of PRS for non-European ancestral populations for CRC. Here we consider two different approaches to PRS development, (1) ancestry-specific PRS using PRS-CSx^15^ based on ancestry-specific GWAS while leveraging cross-ancestry information and (2) single cross-ancestry Asian-European PRS using LDPred2^16^ based on combined meta-analysis summary statistics and LD matrices across Asian and European ancestries. Using independent racially and ethnically diverse datasets, we evaluated the performance of these two PRS and compared them with a genome-wide PRS built using European-only GWAS data^3^ and a PRS based on 204 known CRC loci^17–20^. To facilitate understanding of its clinical utility, we used decision-curve analyses^21^ to assess the standardized net benefit for the model based on family-history and PRS and compared to the family-history-only model, as the latter is currently used to decide at what age screening starts.

## Results

For developing PRSs, we used GWAS summary statistics of 1,020,293 SNPs based on 21,731 cases and 47,444 controls of Asian and 78,473 cases and 107,143 controls of European ancestries. We evaluated the performance of the PRS in independent validation individual-level data sets including 12,025 Asian (2,420 cases; 9,605 controls), 13,823 Black/African-American (1,954 cases; 11,869 controls), 10,378 Latinx/Hispanic (1,682 cases; 8,696 controls) and 118,756 non-Hispanic White (3,651 cases; 115,105 controls) participants. More details about study participant characteristics for training and validation data sets are included in Table 1, Supplemental Table 1, and Supplemental Material and Methods.

**Table 1:**
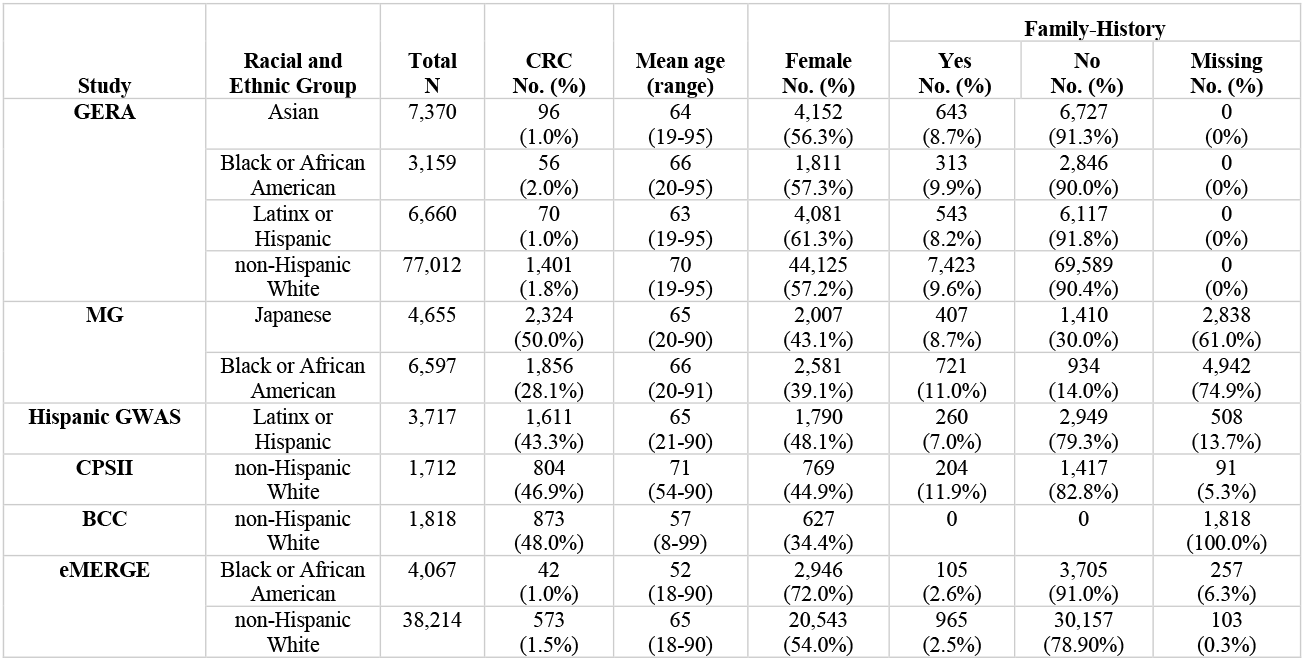
Characteristics of the validation studies

| Study | Racial and Ethnic Group | Total N | CRC No. (%) | Mean age (range) | Female No. (%) | Family-History |  |  |
| --- | --- | --- | --- | --- | --- | --- | --- | --- |
|  |  |  |  |  |  | Yes No. (%) | No No. (%) | Missing No. (%) |
| GERA | Asian | 7,370 | 96 (1.0%) | 64 (19-95) | 4,152 (56.3%) | 643 (8.7%) | 6,727 (91.3%) | 0 (0%) |
|  | Black or African American | 3,159 | 56 (2.0%) | 66 (20-95) | 1,811 (57.3%) | 313 (9.9%) | 2,846 (90.0%) | 0 (0%) |
|  | Latinx or Hispanic | 6,660 | 70 (1.0%) | 63 (19-95) | 4,081 (61.3%) | 543 (8.2%) | 6,117 (91.8%) | 0 (0%) |
|  | non-Hispanic White | 77,012 | 1,401 (1.8%) | 70 (19-95) | 44,125 (57.2%) | 7,423 (9.6%) | 69,589 (90.4%) | 0 (0%) |
| MG | Japanese | 4,655 | 2,324 (50.0%) | 65 (20-90) | 2,007 (43.1%) | 407 (8.7%) | 1,410 (30.0%) | 2,838 (61.0%) |
|  | Black or African American | 6,597 | 1,856 (28.1%) | 66 (20-91) | 2,581 (39.1%) | 721 (11.0%) | 934 (14.0%) | 4,942 (74.9%) |
| Hispanic GWAS | Latinx or Hispanic | 3,717 | 1,611 (43.3%) | 65 (21-90) | 1,790 (48.1%) | 260 (7.0%) | 2,949 (79.3%) | 508 (13.7%) |
| CPSII | non-Hispanic White | 1,712 | 804 (46.9%) | 71 (54-90) | 769 (44.9%) | 204 (11.9%) | 1,417 (82.8%) | 91 (5.3%) |
| BCC | non-Hispanic White | 1,818 | 873 (48.0%) | 57 (8-99) | 627 (34.4%) | 0 | 0 | 1,818 (100.0%) |
| eMERGE | Black or African American | 4,067 | 42 (1.0%) | 52 (18-90) | 2,946 (72.0%) | 105 (2.6%) | 3,705 (91.0%) | 257 (6.3%) |
|  | non-Hispanic White | 38,214 | 573 (1.5%) | 65 (18-90) | 20,543 (54.0%) | 965 (2.5%) | 30,157 (78.90%) | 103 (0.3%) |

### Discriminatory accuracy of Asian-European PRS

The single cross-ancestry Asian-European PRS derived using the combined Asian-European GWAS meta-analysis summary statistics and LD matrices with LDpred2 improved the discriminatory accuracy in the Asian population compared to the European-centric PRS (AUC = 0.63 vs. 0.59, p-value < 4.5e-09, Table 2). It also improved the AUC significantly in the non-Hispanic White population (AUC = 0.65 vs. 0.63, p-value = 6.0e-03). Despite lack of Black/African American and Hispanic individuals in deriving the PRS, the Asian-European PRS improved the AUC for Black/African American (AUC = 0.59 vs. 0.58, p-value = 0.05) and Hispanic individuals (AUC = 0.62 vs. 0.59, p-value = 5.0e-03). The Asian-European PRS improved the AUC in all racial and ethnic groups compared to the known-loci PRS (all p-values < 0.05).

**Table 2:** AUC estimates (95% confidence interval) for European-centric PRS, known loci PRS, PRS-CSx and LDPred2.

| Race and Ethnicity | Cases/ controls | European-centric PRS | Known Loci PRS | PRS-CSx | LDpred2 |
| --- | --- | --- | --- | --- | --- |
| Asian | 2,420/9,605 | 0.59 (0.57-0.60) | 0.60 (0.59-0.62)<br>p-val*: 0.24 | 0.64(0.58-0.69)<br>p-val*: 0.06<br>p-val†: 0.32 | 0.63 (0.62-0.64)<br>p-val*: 4.5e-9<br>p-val†: 1.6e-6<br>p-val**: 0.75 |
| Black or African American | 1,954/11,869 | 0.58 (0.56-0.59) | 0.58 (0.56-0.59)<br>p-val*: 0.92 |  | 0.59 (0.57-0.61)<br>p-val*: 0.05<br>p-val†: 0.01 |
| Latinx or Hispanic | 1,681/8,696 | 0.59 (0.57-0.61) | 0.59 (0.57-0.60)<br>p-val*: 0.76 |  | 0.62 (0.60-0.63)<br>p-val*: 5.0e-3<br>p-val†: 1.0e-3 |
| Non-Hispanic White | 3,651/115,105 | 0.63 (0.62-0.65) | 0.61 (0.60-0.62)<br>p-val*: 9.0e-4 | 0.64(0.62-0.65)<br>p-val* : 0.15<br>p-val† : 1.8e-5 | 0.65 (0.64-0.66)<br>p-val*: 6.0e-3<br>p-val†: 0.11<br>p-val**: 3.0e-3 |
| *p-value comparison of PRS with the European-centric PRS.<br>†p-value comparison of PRS with the known-loci PRS.<br>**p-value comparison of PRS with the PRS-CSx.<br>** All AUC estimates were adjusted for age, sex, and top 4 principal components. |  |  |  |  |  |

The ancestry-specific PRS derived using PRS-CSx improved the discriminatory accuracy in the Asian population compared to the European-centric PRS (AUC = 0.64 vs. 0.59), though not statistically significant with p-value 0.06 (Table 2). The AUC for the ancestry-specific non-Hispanic White-specific PRS was also not statistically different from the European-centric PRS (p-value = 0.15) in the non-Hispanic White population; however, it was significantly higher than the known-loci PRS (p-value = 1.8e-05). The ancestry-specific PRS-CSx is not relevant for Black/African American and Hispanic groups, because there were no GWAS for these groups included in the training datasets.

There was little variation in AUC estimates across studies (Supplemental Table 2). Among these two approaches, the Asian-European PRS using the combined Asian-European summary statistics in LDpred2 had greater discriminatory accuracy than the ancestry-specific non-Hispanic White-specific PRS from PRS-CSx with p-value = 3.0e-03. However, we did not observe statistically significant differences in Asian individuals (p-value = 0.75). Taken together, the single cross-ancestry Asian-European PRS using LDpred2 performs among the best in terms of AUC but with much narrower confidence intervals; hereafter we focus only on the single cross-ancestry Asian-European PRS. The ROC curves for the cross ancestry Asian-European PRS showed a similar pattern to the AUC for Asian, Black/African American, Hispanic, and non-Hispanic White participants (Supplemental Figure 1).

### PRS distribution across racial and ethnic groups

As expected, the PRS distributions varied across the racial and ethnic groups (Figure 1(A) and Supplemental Figure 2). After trans-ancestry correction, the PRS distributions largely overlapped except for the MG-JPN study (Figure 1(B) and Supplemental Figure 3). This may be due to the use of the imputation reference panel of only Asian individuals from the 1000 Genomes Projects for MG-JPN; this differs from all other studies, which used all 1000 Genome Project samples in the reference panel. We thus performed an additional mean adjustment to the PRS for the MG-JPN study. After this adjustment, all PRS distributions overlapped (Figure 1(C)).

**Figure 1:**
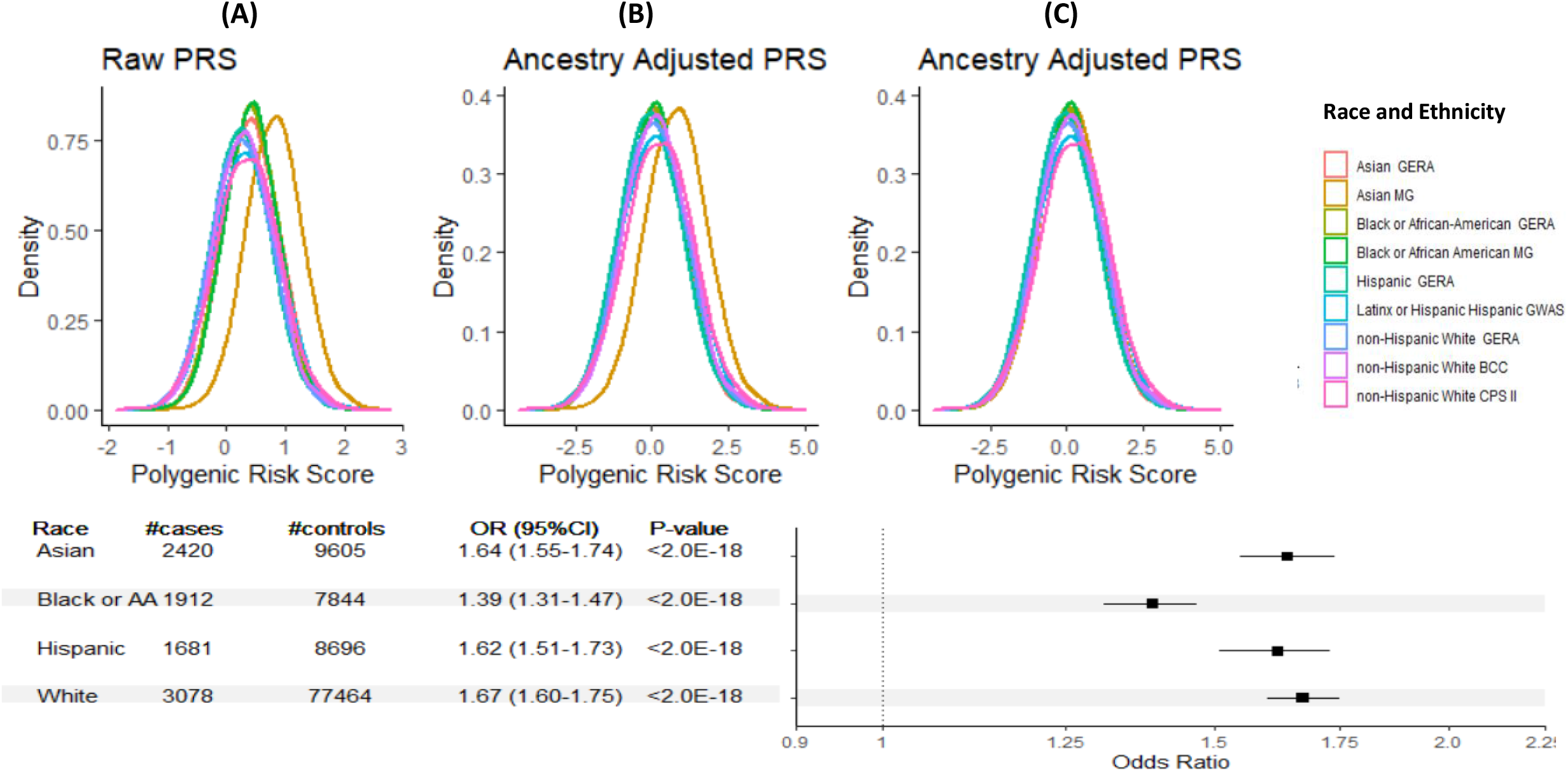
Distribution of PRS: A) PRS distributions varied across racial/ethnic groups, B) PRS distribution after ancestry adjustment, C) Additional mean adjustment for the Asian MG (Minor GWAS Japanese Study) study that has a different imputation panel, and D) forest plot by racial and ethnic group for OR estimates of PRS per SD. PRS is based on single cross-ancestry Asian-European PRS.

Cases had higher mean PRS than controls across all racial and ethnic groups (Supplemental Figure 4). The OR estimates per SD of PRS (95% CI) were 1.64 (1.55-1.74), 1.39 (1.31-1.47), 1.62 (1.51-1.73) and 1.67 (1.60-1.75) for Asian, Black/African American, Latinx/Hispanic, and non-Hispanic White participants, respectively, with p-value < 2.0e-18 for all four groups (Figure 1(D) and Table 3).

**Table 3.**
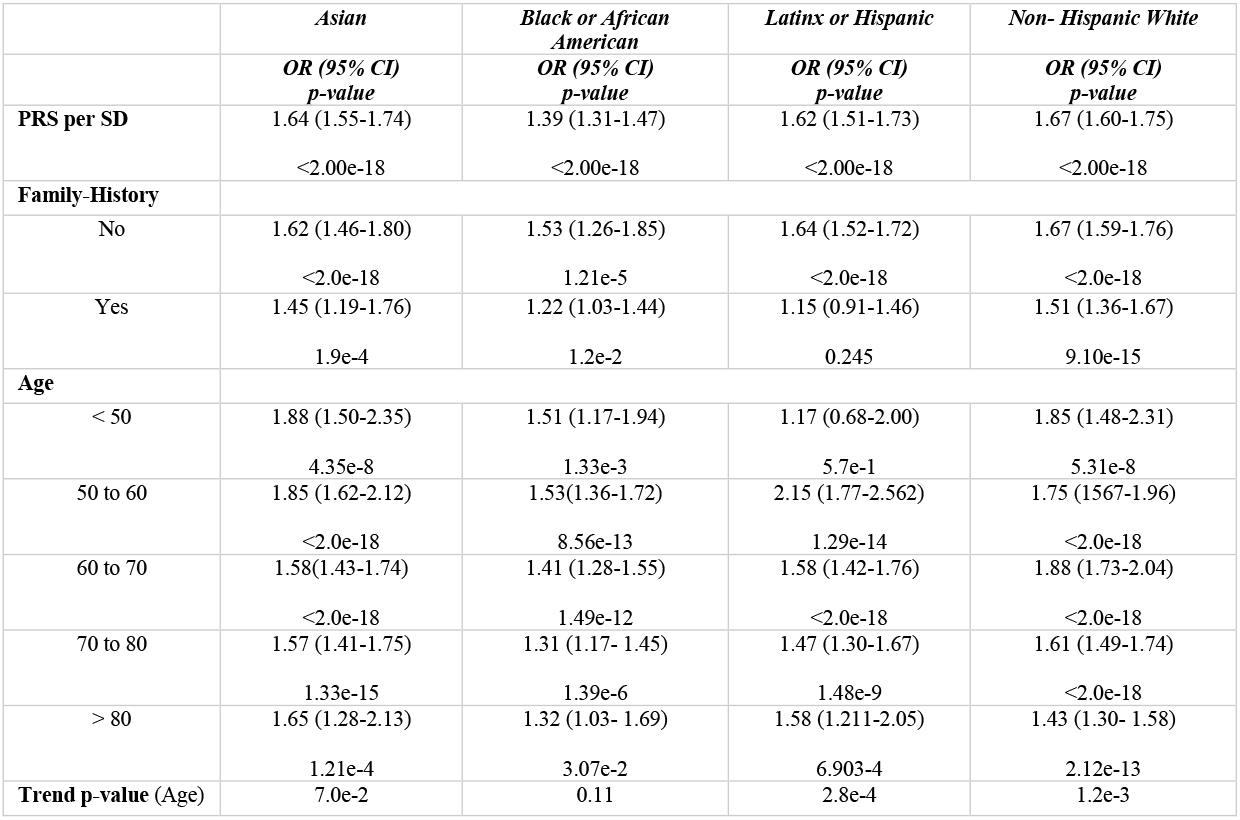
Odds ratios (OR), 95% confidence interval (95% CI) and p-values for PRS per SD and PRS stratified by family-history and age.

Compared to the mean risk, the relative risks of PRS at any given percentile were similar for all racial and ethnic groups except for Black/African American participants for whom it was attenuated (Figure 2). The relative risk at the 90th percentile of the PRS distribution compared to mean was 1.67, 1.44, 1.65, and 1.69 for Asian, Black/African American, Latinx/African American, and non-Hispanic participants, respectively.

**Figure 2:**
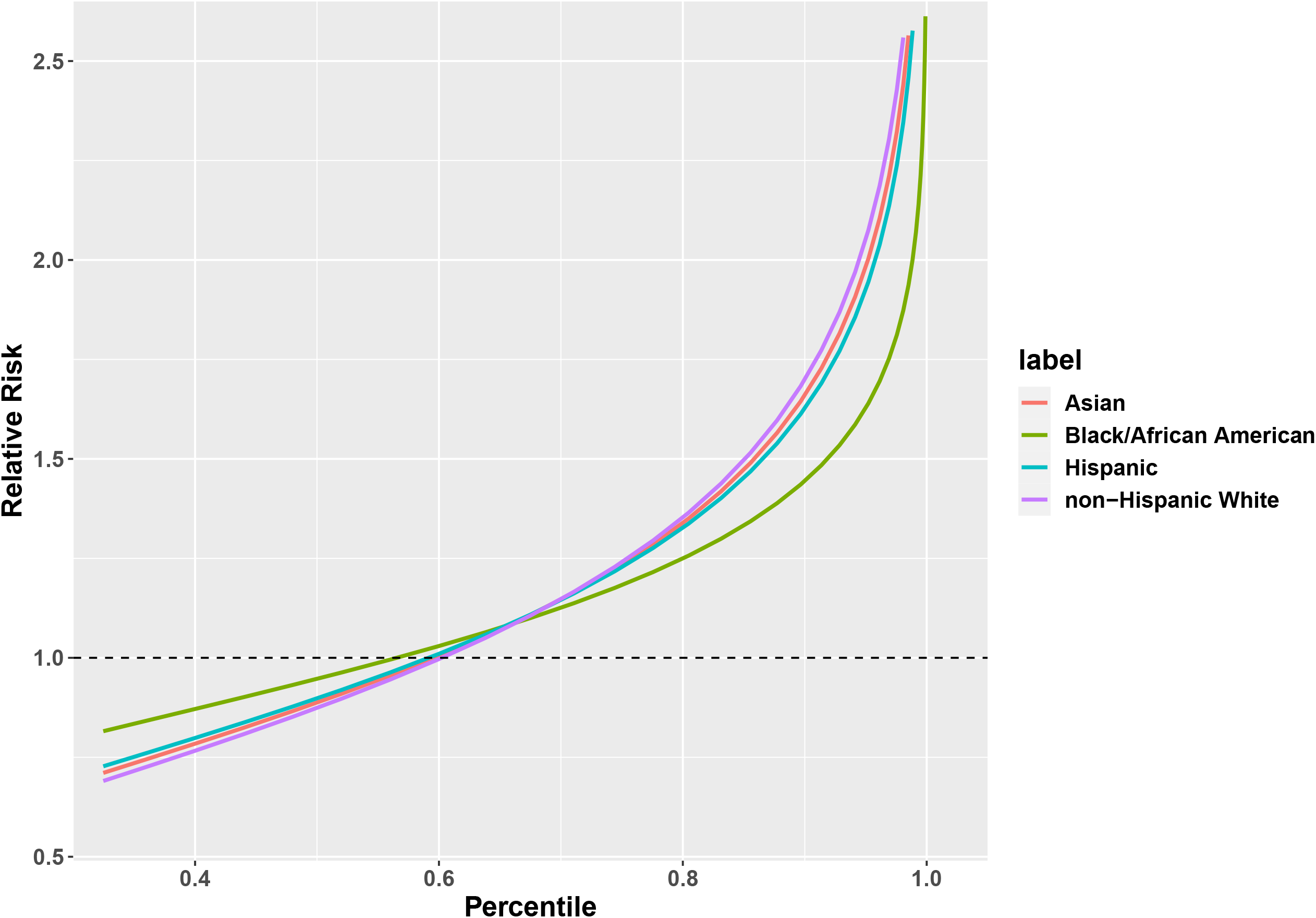
The relative risk of individuals at different percentiles of the single cross-ancestral Asian-European PRS compared to a population average odds ratio, stratified by race and ethnicity

The model-based relative risk was calibrated well across the PRS range in all racial and ethnic groups (Figure 3).

**Figure 3:**
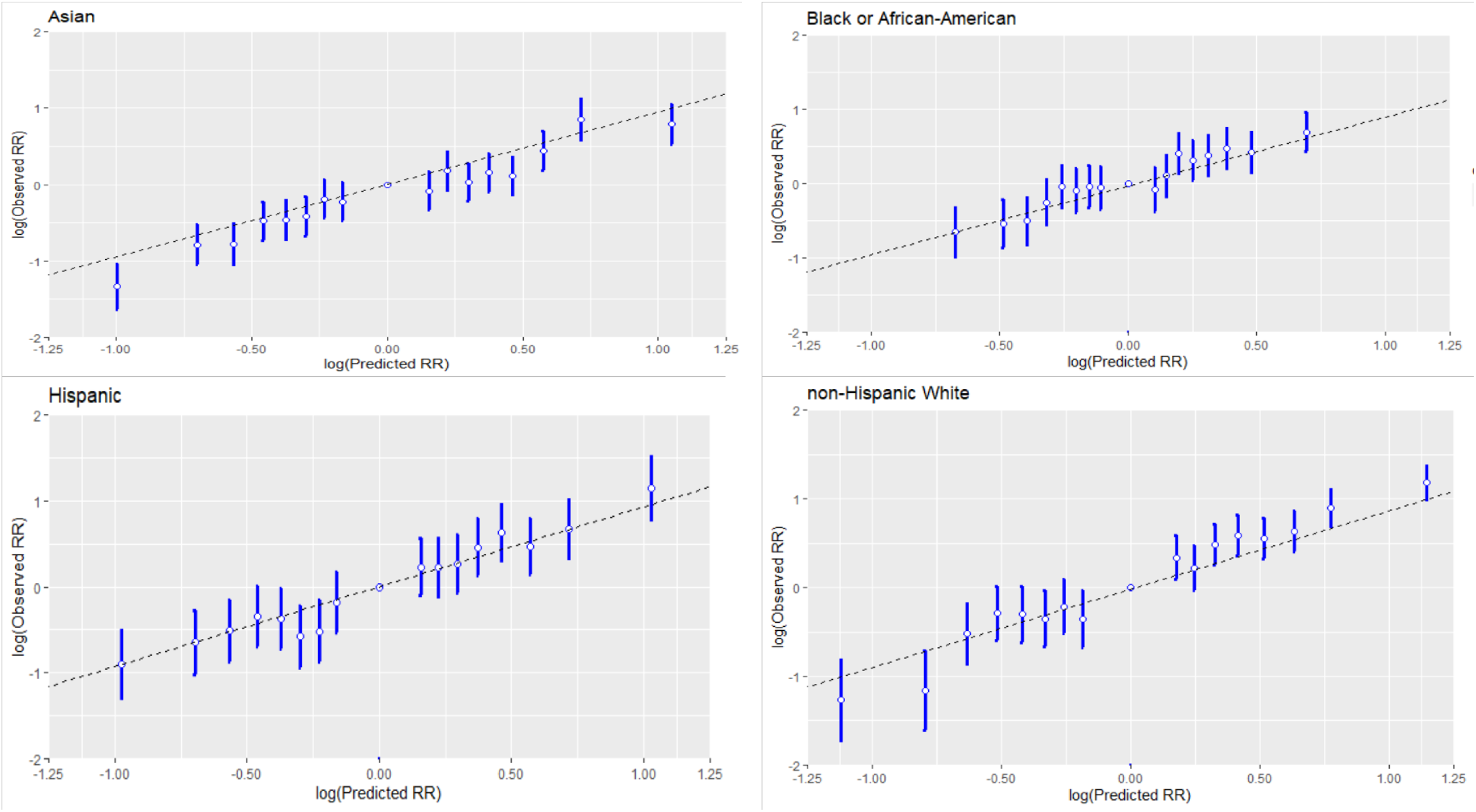
The relative risk calibration of PRS, stratified by race and ethnicity. The x-axis is the log-transformed predicted RR values and the y-axis is the log-transformed observed RR with the middle bin (40-60) as the reference group.

### Odds ratios (ORs) for PRS stratified by family-history and age

Across all racial and ethnic groups, the ORs for the PRS were higher in those without a family-history than those with a family-history with p-values 0.21, 0.01, 3.0e-3, and 0.11 for Asian, Black/African American, Latinx/Hispanic, and non-Hispanic White participants respectively (Table 3). The estimates were consistent across studies (Supplemental Table 3).

The strength of association estimates for PRS in relation to CRC decreased over strata of increased age in each racial and ethnic group with trend test p-values of 0.07, 0.11, 2.8e-4, and 1.2e-03 for Asian, Black/African American, Latinx/Hispanic, and non-Hispanic White, participants, respectively. The ORs, 95% CI and trend p-value for each racial and ethnic group are given in Table 3. The estimates were consistent across studies (Supplemental Table 3).

### Clinical utility for model based on PRS and family-history

We calculated the standardized net benefit (sNB) to assess the clinical utility of using a model based on PRS and family-history to recommend an intervention (such as screening) for participants < 50 years of age. We used the average 10-year risk of developing CRC at age 45 as the risk threshold, because the current CRC-screening guidelines recommend that an average-risk individual start screening at age 45 years old. Using the GERA cohort, we estimated the 10-year risk to be 0.29% across all racial and ethnic groups. At this risk threshold, the risk model based on PRS, and family-history achieved 37.3% (95% CI:23.8%-50.8%) of the maximum possible achievable utility. This was greater than the model based on family-history alone (sNB =21.7%, 95% CI: 12.4%-33%, p-value 0.02) and hypothetically intervening on all or no people (Figure 4), a pattern that generally holds for each racial and ethnic group (Supplemental Figure 5).

**Figure 4:**
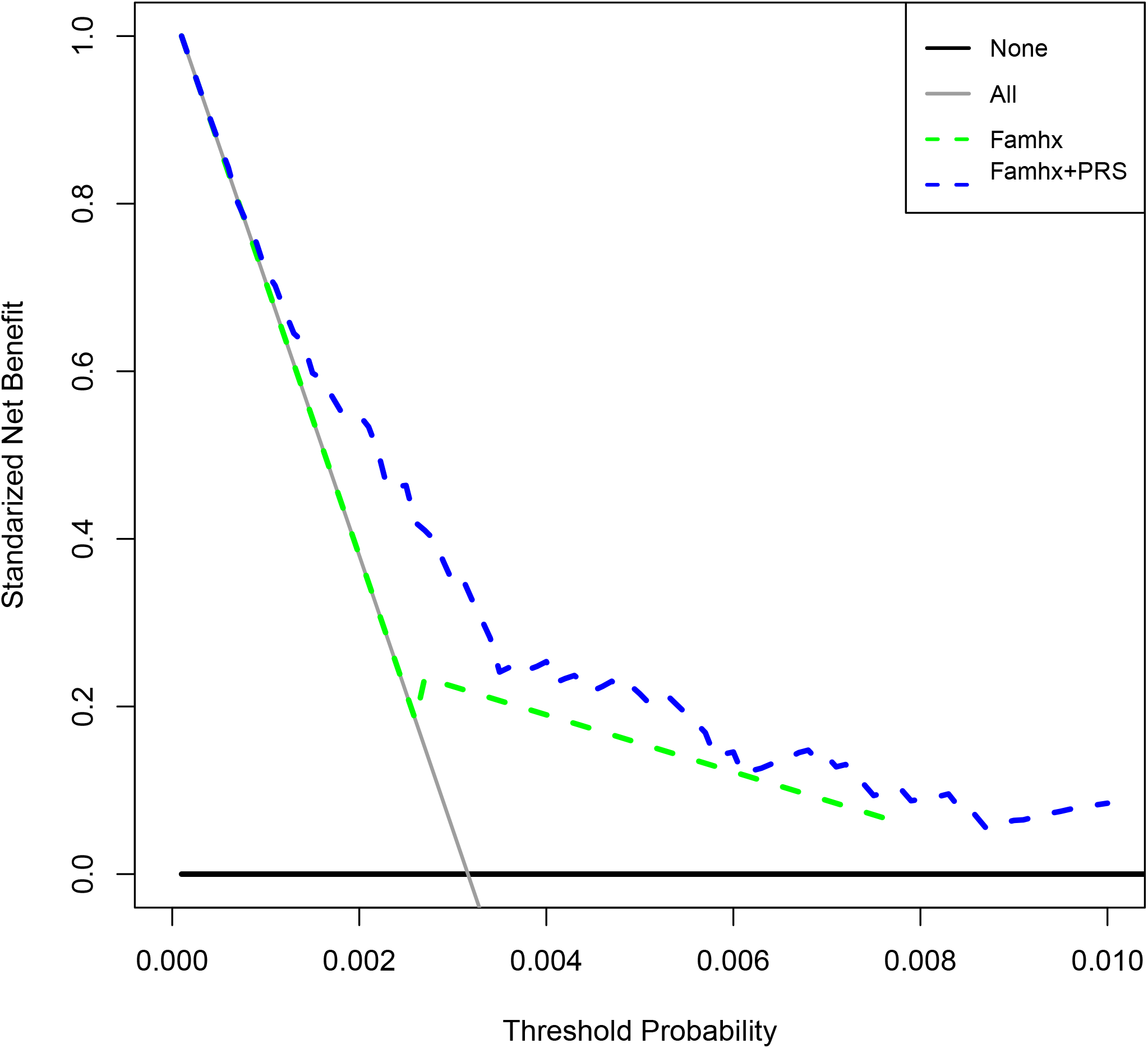
Decision-curves for the model based on family-history and PRS, model based on family-history alone, intervene for all, and intervene for none, in GERA study participants with age 40-49 (22,628 participants with 149 cases). X-axis is the range of threshold probabilities (CRC-risk). The legends of the lines are solid black: intervene for none (none); solid gray: intervene for all (All); dash green: model based on family-history (Famhx); dash blue: model based on family-history and PRS (Famhx + PRS).

We observed a similar pattern for participants between the ages of 50 and 60 years (Supplemental Figure 6). We also used the 10-year risk 0.39% at age 50 and 0.49% at 55 years as the risk thresholds. The risk model based on PRS, and family history achieved greater sNB (sNB = 24.8% and 21.6%, respectively) than the model based on family history-alone (sNB = 19.3% and 15.9%, respectively).

## Discussion

Using large-scale Asian and European GWAS data, we demonstrate that combining Asian and European summary statistics in deriving PRS led to statistically significant improvement in discriminatory accuracy across Asian, Black/African American, Latinx/Hispanic and non-Hispanic White groups, although the improvement was less marked in Latinx/Hispanic and Black/African American participants. We further show that across all groups, the PRS has stronger associations with CRC-risk in younger individuals and in those without a family-history of CRC, which will likely increase the possible clinical utility of the PRS given the rising young-onset CRC incidence rates in recent decades, mostly in individuals without a known family-history. This is supported by our decision-curve analysis demonstrating that adding PRS improves the maximum achievable clinical utility over the model based on family-history only for ages 40-60 years.

A challenging factor of moving PRS to clinical implementation is ensuring that the PRS is equally applicable to individuals across all racial and ethnic groups to prevent an increase in health disparities. Relevant to this objective, we evaluated two broad categories of approaches (ancestry-specific PRS while leveraging cross ancestry information and single cross-ancestry PRS based on the combined cross-ancestry GWAS) for improving the prediction in under-represented groups, and our observation of the performance of these approaches could be generalized to other traits besides CRC. We found that both approaches performed similarly in Asian and non-Hispanic European individuals. Further, the cross-ancestry Asian-European PRS also improved risk prediction in Black/African American and Hispanic groups, though to a lesser extent. We also show that we can correct this raw PRS for genetic ancestry and create a common distribution that can be used across racial and ethnic groups, avoiding the potential difficulty of using ancestry-specific PRS in admixed populations. Accordingly, our cross-ancestry Asian-European PRS has the potential to reduce health disparities between non-European ancestry populations and the European ancestry population.

As there is growing interest in clinical use of PRS, it is important to point out that the purpose of PRS is not to identify CRC, but rather stratify individuals into different risk strata for which different levels of cancer preventive interventions may be devised.^22,23^ Their performance should thus be compared with risk factors currently used for risk stratification such as family-history in terms of cost effectiveness. In this paper, we performed a decision-curve analysis that has been used in cancer research for assessing the potential population impact of incorporating a risk prediction model into clinical practice^24–26^. The risk model that incorporates both the PRS and family-history achieves 37.3% of the maximum possible achievable utility for those 40-49 years old, significantly greater than 21.7% under the family-history-only model. Recently the US Preventive Services Task Force recommended lowering the age at screening initiation to 45 years for individuals at average risk^27^. However, given the substantial burden of additional approximately 22 million people becoming eligible for screening and the fact that CRC remains a rare event in younger individuals, there has been critique of the universal change to the initial screening age that, instead, emphasizes the importance of targeted screening based on an individual’s risk factors^28–30^. The results from the decision-curve analysis suggest that there is clinical utility to adding a PRS to the family-history-only model in risk stratification for CRC prevention. To fully evaluate the effectiveness of including PRS as part of risk stratification, a full decision analytic modeling that incorporates other aspects such as different screening methods, implementation factors, behavioral factors, and corresponding costs are warranted^31^.

Recent efforts^32,33^ in clinical implementation of PRS shows the potential of PRS to effectively stratify the risk of diseases development and guide screening. BOADICEA v5 (as implemented in the CanRisk tool)^32^ already implements a 313-variant PRS of breast cancer and currently supports hundreds of thousands of women, doctors, and genetic counselors annually in >90 countries making treatment decisions. PRS-guided mammographic screening is also being tested in the WISDOM and PERSPECTIVE I&I studies^33^. GenoVA Study ^34^ is a clinical trial in which patients and their primary care physicians receive a clinical PRS laboratory report on five diseases including CRC. MyOme implements a cross-ancestry risk score for breast cancer risk stratification ^35^. As CRC has an effective screening intervention, it would be of great interest to explore implementation of PRS for guiding personal screening recommendations.

This study has several strengths. We brought together most of the globally available GWAS of CRC for Asian and European ancestry populations as our training data, which is an important factor for the improved performance of the proposed PRS. Further, we used multiple independent evaluation data sets that were not part of our training data nor GWAS discovery, providing an unbiased evaluation of the developed models. Moreover, the single cross-ancestral PRS derived in this study makes it easy to implement in any admixed population.

The results of this investigation should be interpreted in the context of its limitations. The discriminatory accuracy remains lower in Latinx/Hispanic and particularly in Black/African American individuals due to their limited sample sizes in training data. Future studies more inclusive of these individuals are warranted for deriving PRS to enhance the discriminatory accuracy. Furthermore, we have not been able to evaluate the performance of these models in other racial and ethnic groups, including Alaskan Native, Native American and Pacific Islander individuals. Lastly, we expect to further improve risk prediction by combining the PRS with non-genetic risk factors such as obesity, diet, and aspirin use, as previously shown^23,36^.

Advances in PRS development have promoted the use of PRS-enhanced models to determine and stratify disease risk, which could improve disease prevention and management through screening and early detection. Our cross-ancestry Asian-European PRS, built upon data on both Asian and European ancestry individuals, improves the PRS performance in Asian, Black/African, and Latinx/Hispanic individuals considerably. Combining PRS and other CRC-associated risk factors such as lifestyle/environmental risk factors and high penetrance genes will likely further improve the prediction performance^36^. We anticipate that the continuous expansion of PRS development and validation to include more diverse populations and prospective evaluation of PRS-enhanced risk prediction model in clinical trials along with decreasing genotyping cost and adaptation of health care systems to accommodate genetic data and prediction algorithm will bring closer the implementation of PRS in clinical practice.

## Methods and Materials

### Training Data Sets

To develop polygenic risk scores (PRS) across population, we used the genome-wide association study (GWAS) summary statistics of 1,020,293 SNPs based on 78,473 cases and 107,143 controls of European (EUR) and 21,731 cases and 47,444 controls of Asian ancestries from previously published GWAS (Supplemental Table 1)^17–19^. For this we group participants into analytical units by study or genotyping platform as consistent with the original reports^17–20,37,38^. Ancestry was determined by the genetic principal component analysis. Studies that contributed to more than one prior genome-wide association analyses were analyzed only once. In total, there were 31 analytical units (17 from EUR descent populations and 14 from Asian descent populations), totaling 100,204 CRC cases and 154,587 controls. Comprehensive details on the participants, genotyping and standard quality control (QC) procedures have been previously reported and are also summarized in Supplemental Table 1. All study protocols were approved by the relevant Institutional Review Boards, and informed consent was obtained from all study participants in accordance with the Helsinki accord.

### Independent Validation Data Sets

We evaluated the performance of each of the developed PRS in the Genetic Epidemiology Research on Adult Health and Aging Cohort (GERA) cohort; Minority GWAS Japanese study (MG-JPN)^39^; Minority GWAS African American study (MG-AA)^40^; Hispanic Colorectal Cancer Study (HCCS)^41^; Multiethnic Cohort study (MEC); Cancer Prevention Study II (CPSII)^42^; Basque-colon cohort (BCC); and Electronic Medical Records and Genomics (eMERGE) study. Racial and ethnic identification in these studies were self-reported. In total, there were 12,025 Asian (2,420 cases; 9,605 controls), 13,823 Black/African-American (1,954 cases; 11,869 controls), 10,378 Latinx/Hispanic (1,682 cases; 8,696 controls) and 118,756 non-Hispanic White (3,651 cases; 115,105 controls) participants. None of these samples was included in the training data sets for model building. More details about study participant characteristics are included in Table 1.

CRC status (Yes/No) was determined from cancer-registry data. Family-history of CRC (>=1 first-degree relatives with CRC), was ascertained through baseline study questionnaire or electronic medical records at study entry.

### Approaches for Deriving PRS

We compared two different approaches for PRS development using 1) ancestry-specific PRS using PRS-CSx that integrates genome-wide Asian and European summary statistics and LD matrices; 2) single cross-ancestry PRS using LDpred2 that combine genome-wide Asian and European summary statistics and a weighted LD matrix with weight defined as the proportion of participants from each ancestry in the summary statistics. Figure 5 depicts the summary of these PRS derivations.

**Figure 5:**
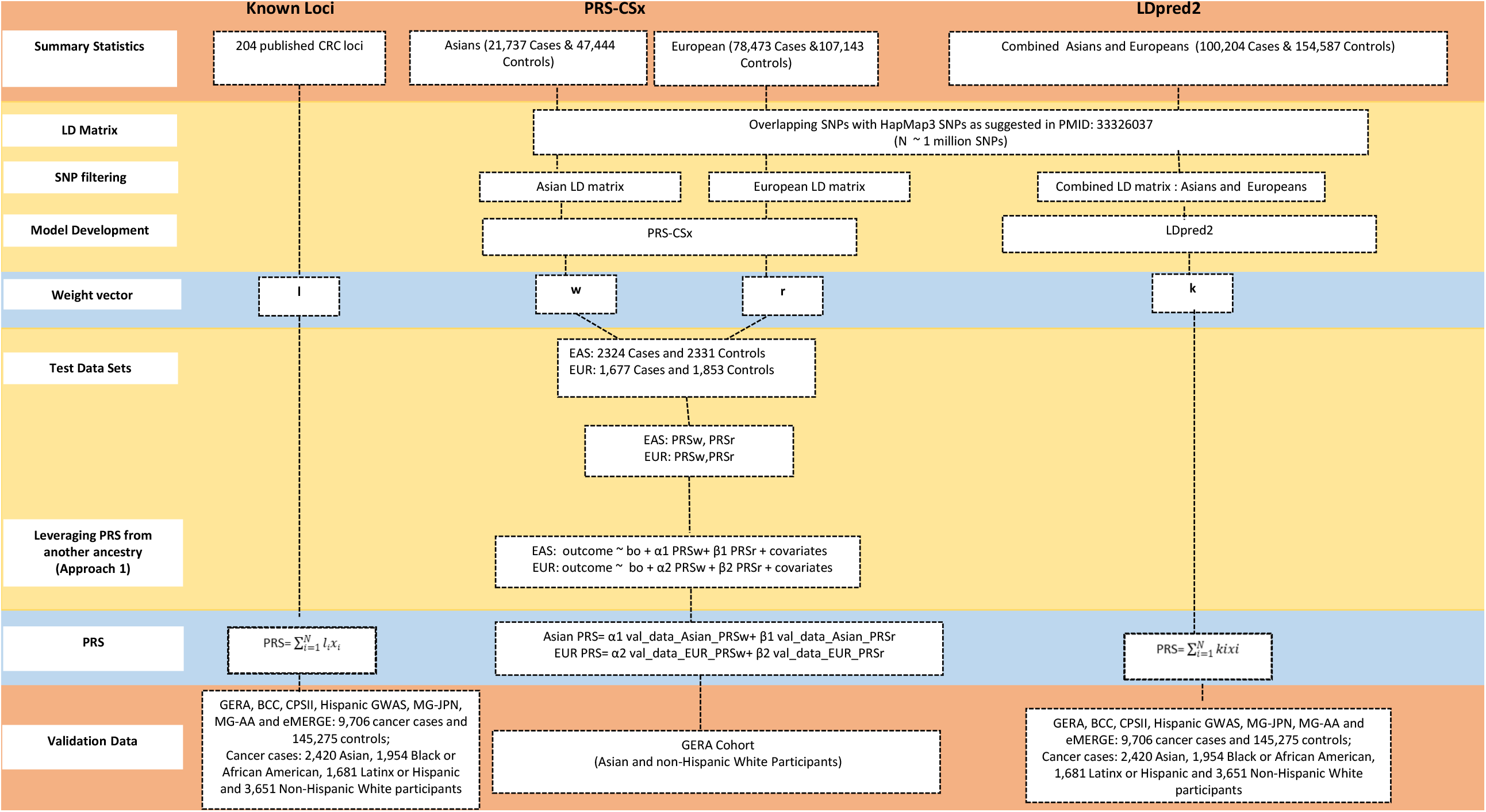
Different approaches for deriving polygenic risk scores (PRS) for colorectal cancer

PRS-CSx^15^ derives ancestry-specific PRS while leveraging GWAS summary statistics from other ancestral groups. We first obtained ancestry-specific PRS using ancestry-specific GWAS summary statistics and LD matrix for Asian and non-Hispanic White participants based on ∼1M genome-wide SNPs, respectively, while leveraging GWAS from the other ancestral group. We denoted these PRS by PRS_Asian_ and PRS_European_, respectively. We then improved ancestry-specific PRS by taking a weighted sum of these PRSs to predict CRC of respective ancestral group. To derive PRS for the Asian population, we calculated a weighted sum of PRS_Asian_ and PRS_European_ (α_1_ PRS_European_+β_1_ PRS_Asian_) and obtained α_1_ and β_1_ from a logistic regression model using the MG-JPN study. Similarly, to derive PRS for the European population, we calculated a weighted sum of PRS_Asian_ and PRS_European_ (α_2_ PRS_European_+β_2_ PRS_Asian_), where α_2_ and β_2_ were obtained based on the pooled BCC and CPSII studies.

To derive the single cross-ancestry PRS using LDpred2^16^, we combined the summary statistics from the Asian and European GWAS using the inverse variance weighted estimator^43^ and combined the LD matrices, as the weighted sum of the Asian and European-specific LD matrices with the weights proportional to the sample sizes of the Asian and European individuals in the combined summary statistics.

We compared ancestry-specific and single cross-ancestry PRS from PRS-CSx and LDpred2 with a previously published European-centric genome-wide PRS^3^ and a known-loci PRS consisting of 204 independently CRC-associated variants based on GWAS of European and Asian ancestries^17–20^ (Supplemental Table 4).

### Evaluation of Model Performance

#### The Area Under the Receiver Operating Characteristics curve (AUC)

We evaluated the predictive performance of the PRS by the area under the receiver operating characteristics curve (AUC) in each of the racial and ethnic groups^44^. We calculated the adjusted AUC of PRS for each study using the ROCt R package^45^, adjusting for covariates age, sex and four PCs. We emphasize that the AUC estimate was for PRS only and the covariates were not part of prediction along PRS. These covariates were included as potential confounders. We then combined the AUC estimates of PRS across studies for each ancestry using the inverse variance weighted estimator.

We obtained the bootstrapped-based standard error (se), 95% confidence intervals (CI) (1.96* se) and two-sided p-values for comparisons across various subgroups using 500 bootstrap samples.

#### Ancestry Adjustment of PRS Distribution

As the PRS distributions were different across racial and ethnic groups due to different allele frequencies, we used a modified trans-ancestry adjustment of PRS to align the PRS distributions^46^. We used the 1000 Genome dataset to estimate the ancestry adjustment following the approach in Khera *et al*. (2019)^46^. Specifically, we derived principal components (PCs) based on 343,662 ancestry informative SNPs with little overlapped (0.3%) with SNPs used in PRS development. To correct for the mean and variance differences between ancestry groups, we fit two linear regression models to predict the mean and variance of PRS based on the first four PCs. To correct for the raw PRS distribution in our data set, we first calculated the PCs using the same loadings for the top 4 PCs from the 1000 Genome data set. We then obtained the ancestry-adjusted PRS for each individual by subtracting the predicted mean based on the 4 PCs from the individual’s raw PRS and then divided it by the predicted standard deviation based on the 4 PCs. Additional adjustments are needed for data sets with different imputation panels. The ancestry adjusted PRS is computed as given below:

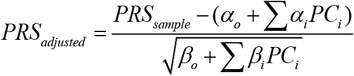

#### Odds Ratio (OR) Estimates

We estimated the OR and 95% CI of CRC-risk associated per SD change in PRS by logistic regression model, overall and stratified by family history and age. For each racial and ethnic group, we estimated the AUC and OR by study and combined the estimates using the inverse variance weighted estimator. In addition, we estimated OR stratified by family history of 1st degree relative with CRC (yes, no) and age (<50, 50-59, 60-69, 70-79, and >80). All analyses were adjusted for age, sex, and top 4 principal components of ancestry.

#### Relative Risk Calibration of PRS

We binned PRS into 5% strata and defined the reference group as PRS in the 40%-60% stratum. The expected OR for a PRS stratum is the ratio of the within-stratum geometric average of individuals’ model-based OR, defined as exponent of individuals’ PRS times log (OR), between that stratum and the reference stratum. We estimated the observed OR estimates and its 95% CI by fitting a logistic regression model with CRC disease status as outcome and a binary variable with 1 indicating a specific stratum and 0 indicating the reference stratum, adjusting for age, sex, and first four principal components.

#### Decision Curve Analysis

The decision-curve analysis was performed by calculating the standardized net benefit (sNB), defined as the net benefit divided by the maximum possible net benefit^21^, to assess the potential clinical impact of the risk prediction models on recommended interventions (i.e., screening). For a given risk threshold, the NB was defined as sensitivity × p – (1 – specificity) × (1 – p) × w, where w was the odds at the threshold, sensitivity was the proportion of cases above the risk threshold based on the model, specificity was the proportion of controls below the risk threshold based on the model, and p was the disease probability at the landmark time. As it was difficult to interpret NB itself, we followed the approach proposed by Kerr *et al*.^21^ to calculate sNB, i.e., dividing NB by the maximum NB, which is achieved when sensitivity = 1 and specificity = 1. Hence, the sNB was equal to sensitivity – (1 – specificity) × (1 – p)/p × w. It provided some sense of magnitude of sNB on a percent scale and was interpreted as the relative utility that has maximum value of 1. For example, if sNB = 0.4, it means that the risk model achieves 40% of the maximum possible achievable utility.

We compared the model based on PRS and family history with the model based on family history alone, as well as two hypothetical extreme scenarios: intervention (e.g., screening) for all and intervention for none. We calculated the sNB under the competing risks framework^47^, where the observational time is the minimum of time to CRC, time to death, and time at last observation, and the disease status is 1 if the study participant had CRC, 2 if the participant died (competing event), and 0 otherwise. We used GERA to calculate sNB, as this was the only cohort study among our independent validation data sets. We plotted decision-curves of sNB at the 10-year landmark time vs. risk threshold for age at study entry 40-49 and 50-59 years old, because average-risk individuals in these age groups are recommended to start CRC screening.

We performed the analyses using R packages^24,45,48–50^. A two-sided p-value < 0.05 is considered statistically significant.

## Data Availability

No primary data were collected in the present study. The GWAS summary statistics is available from GWAS catalog (https://www.ebi.ac.uk/gwas/) with accession number GCST90129505. PRS SNP weights will be deposited in the PGS catalogue repository (https://www.pgscatalog.org/).

