## Supplemental Text for "Combining Asian-European Genome-Wide Association Studies of Colorectal Cancer Improves Risk Prediction Across Race and Ethnicity"

**Supplementary Methods and Results**

***Test and Validation Data Sets***

***The Minority GWAS Japanese study (MG-JPN)***

The Japanese samples (n =4,655) were identified from the Multiethnic Cohort study (MEC), the Colorectal Cancer Family Registry (CCFR), the Japan Public Health Center-based Prospective study (JPHC) and three case–control studies conducted in Hawaii (CR2&3) and in Fukuoka and Nagano, Japan. Comprehensive details on the participants subjects, genotyping and standard quality control (QC) procedures have been previously reported in[^1^](https://sciwheel.com/work/citation?ids=1638967&pre=&suf=&sa=0&dbf=0) and are summarized in **Table 1.**

***The Minority GWAS African American study (MG-AA)***

The African American samples (n = 6,597) were identified from the MEC, CCFR, the Southern Community Cohort Study (SCCS), the MD Anderson Cancer Center, the University of North Carolina CanCORS study (UNC-CanCORS) and Rectal Cancer Study (UNC-Rectal), and from the Prostate, Lung, Colorectal and Ovarian Cancer Screening (PLCO) Trial. The details on the participants, genotyping, QC procedures were published in [^1^](https://sciwheel.com/work/citation?ids=1638967&pre=&suf=&sa=0&dbf=0) and are summarized in Table 1.

***The Hispanic Colorectal Cancer Study (HCCS)***

The Hispanic Colorectal Cancer Study (HCCS) is a population-based study of individuals self-identified as Hispanic with a diagnosis of CRC. Cases were identified from the California Cancer Registry or directly from local hospitals in the Los Angeles region [LAC + USC County Hospital and University of Southern California (USC) Norris Comprehensive Cancer Center]. All men and women over 21 years of age with a first-time diagnosis of CRC (ICD-O-3 codes: C18–C21) after January 1, 2008, were eligible for participation. Risk factor/dietary questionnaires, pathology reports and saliva samples (for genotyping) were collected using methodologies developed in the Colon Cancer Family Registry and the Multiethnic Cohort (MEC). The present study includes 950 cases recruited into the HCCS who were born in Mexico (42.3%), the USA (31.4%), Central/South America (16.6%), Cuba or the Caribbean Islands (1.8%) or Europe (0.4%). This study was approved by the USC Institutional Review Board and the California Committee for the Protection of Human Subjects, and all participants provided written informed consent. The detailed descriptions of the participants, genotyping, QC procedure were given in [^2^](https://sciwheel.com/work/citation?ids=1804225&pre=&suf=&sa=0&dbf=0) and are summarized in Table 1.

***Multiethnic Cohort Sstudy (MEC)***

The Multiethnic Cohort Study (MEC) [^2^](https://sciwheel.com/work/citation?ids=1804225&pre=&suf=&sa=0&dbf=0) is a large prospective cohort study that includes people from various ethnic groups, including Hispanic/Latino (HL) primarily from California and mainly, Los Angeles. Between 1993 and 1996, participants returned a self-administered baseline questionnaire that obtained general demographic, medical and risk factor information. The MEC used state driver’s license files as the primary source to identify study participants in California. Surnames were used to identify HL individuals because race/ethnicity was not available in driver’s license files.

In the cohort, incident cancer cases were identified annually through cohort linkage to population-based Surveillance, Epidemiology and End Results cancer registries in Los Angeles County as well as to the California State cancer registry in the same manner as in the HCCS. All men and women over age 21 with a first-time diagnosis of CRC (ICD-O-3 codes: C18–C21) were included as eligible cases. The current study used questionnaire data and DNA samples derived from whole blood or buccal cells for 661 HL prevalent or incident CRC cases born in the USA (57.8%), Mexico (27.7%), Central/South America (9.7%), Cuba or the Caribbean Islands (4.4%), or Europe (0.2%). Individuals without a diagnosis of CRC were used as controls (n=2,106). All MEC controls self-reported being born in the USA (52.2%), Mexico (34.3%), or Central/South America (13.2%). This study includes 661 cases and 2,106 controls.

This study was approved by the University of Southern California and the University of Hawaii Institutional Review Boards, and all participants provided informed consent.

***Cancer Prevention Study II (CPS II)***

The CPS II Nutrition cohort is a prospective study of cancer incidence and mortality in the United States, established in 1992 and described in detail elsewhere [^3^](https://sciwheel.com/work/citation?ids=2831054&pre=&suf=&sa=0&dbf=0). At enrolment, participants completed a mailed self-administered questionnaire including information on demographic, medical, diet, and lifestyle factors. Follow-up questionnaires to update exposure information and to ascertain newly diagnosed cancers were sent biennially starting in 1997. Reported cancers were verified through medical records, state cancer registry linkage, or death certificates. The Emory University Institutional Review Board approved all aspects of the CPS II Nutrition Cohort.

Two independent sets of CPS II cases and matched controls were included in the training data set described above. A third independent set of 804 cases and 908 and sex- and age-matched controls were included in this validation effort.

***Basque-colon cohort***

Basque-colon cohort included 873 participants with colon cancer, biobanked between 2009-2019.  Participants were diagnosed and/or treated according to usual clinical practice in the Donostia University Hospital, San Sebastian, Spain, including follow-up to this date. Controls (n = 945) came from the blood donor biobank of the same geographical area.

***Genetic Epidemiology Research on Adult Health and Aging (GERA) Cohort***

The Genetic Epidemiology Research on Adult Health and Aging (GERA) resource is a cohort of more than 100,000 participants from Kaiser Permanente Medical Care Plan, Northern California Region (KPNC), Research Program on Genes, Environment and Health (RPGEH). Genome-wide genotyping was targeted for this cohort to enable large-scale genome-wide association studies by linkage to comprehensive longitudinal clinical data derived from extensive KPNC electronic health record databases. The cohort is multi-ethnic, with ∼20% minority representation (African American, East and Southeast Asian, and Hispanic/Latinx or mixed), and the remaining 80% non-Hispanic white. For this project, four ancestry-specific arrays were designed based on the Affymetrix Axiom Genotyping System. Imputation was performed using 1000 Genomes data on an array-wise basis.

Construction of this cohort began in 2007, when a six-page survey was mailed to 1.9 million individuals, age 18 and older, who had been previously enrolled health plan members for at least two years. This survey was designed to ascertain data on demographic and lifestyle characteristics, including race/ethnicity, education, income, marital status, and family history for 35 selected conditions, diet and physical activity, smoking and alcohol consumption, and reproductive history and health. In July 2008, approximately 400,000 survey respondents were asked to sign a consent form authorizing broad use of their survey data, longitudinal electronic health record data, and biospecimens in conducting research on genetic and environmental factors associated with health and disease. Those who provided consent were mailed saliva DNA collection (Oragene) kits. In 2009, over 40,000 men ages 45 to 69 years, who were KPNC health plan members and had enrolled in the California Men’s Health Study (CMHS) in 2002–2003, were similarly asked to provide saliva samples and added to expand the GERA cohort. At CMHS enrollment, men completed mailed surveys to ascertain data on demographic and lifestyle factors, akin to that of GERA.

In total, 110,266 consenting participants who provided saliva samples were selected for genotyping. All racial and ethnic minority participants with saliva samples (n = 20,935, 19%) were included to maximize diversity. Four custom Affymetrix Axiom arrays were designed for genotyping, one for each major ancestral group represented in the GERA cohort: European, African, East Asian, and Latinx. As detailed elsewhere [^4^](https://sciwheel.com/work/citation?ids=886537&pre=&suf=&sa=0&dbf=0), the selected number of SNPs and SNP content varied by array in order to maximize coverage of the whole genome, along with common and low frequency SNPs specific to race/ethnicity and known SNPs associated with disease phenotypes. Details on the calling and quality control have been described previously [^5^](https://sciwheel.com/work/citation?ids=1259401&pre=&suf=&sa=0&dbf=0).

A personal history of cancer was determined from cancer registry and electronic health record data, medical record coding and electronic pathology report coding. A family history of CRC was ascertained through integrating data from baseline surveys and electronic health records (i.e., diagnosis codes, family history documentation). All study participants provided written informed consent, and the study was approved by the KPNC Institutional Review Board. As a cohort unselected on any disease phenotype, GERA participants were not asked to engage in specific medical or screening tests for research purposes. Therefore, although the majority (69%) of GERA participants were age 55 and older at baseline, most, but not all, have undergone screening for colorectal cancer, either by fecal immunochemical testing (FIT) or endoscopy (sigmoidoscopy or colonoscopy). At their baseline questionnaire, 70% were up to date for colorectal cancer screening.

Colorectal cancer status was determined for study participants from their initiation of KPNC health plan membership by linkage to the KPNC Cancer Registry, which captures data on all participants with newly diagnosed cancers at KPNC facilities that adheres to the National Cancer Institute’s Surveillance, Epidemiology, and End Results (SEER) Program standards. The observed time was defined from the age of initial KPNC enrollment to the earliest of age at CRC diagnosis, death or end of follow-up (until December 31, 2016), truncated age to 95, and the CRC incidence is measured from 2007 to 2016. While stratifying participants based on family history, we classified them as yes for those who have positive family history of CRC and no with those who have no information on family history. After quality control, we included 77,012 non-Hispanic White (1,401 cases; 75,611 controls), 7,370 Asian (96 cases; 7,274 controls), 3,159 African American (56 cases; 3,103 controls) and 6,660 Latinx/Hispanic (70 cases; 6,590 controls) participants. CRC status was determined from cancer-registry data.

***Electronic Medical Records and Genomics (eMERGE)***

The eMERGE network has been developing a unified genome‐wide single‐nucleotide variant (SNV) genotype array‐based association platform for analysis of electronic medical record (EMR)‐derived phenotypes for approximately 10 years[^6–8^](https://sciwheel.com/work/citation?ids=7571052,378397,111800&pre=&pre=&pre=&suf=&suf=&suf=&sa=0,0,0&dbf=0&dbf=0&dbf=0). In the first phase, eMERGE 1, discovery efforts were based on the Illumina 660k genotype array with ~20,000 participants being enrolled through five medical centers. In eMERGE 2 ~30,000 more individuals with high‐density genotype data were ascertained resulting in analyses with ~50,000 individuals. In eMERGE 3, genotype and clinical EMR data of ~33,000 additional participants have been added to the resources available for analysis. The case control status for CRC was defined by an algorithm based on ICD9 codes 153 -153.9, 154 - 154.2,154.8 and ICD10 codes C18-C18.9, C19, C20, C21-C21.2, C21.8 (<https://phekb.org/phenotype/colorectal-cancer-crc>). After removal of low‐call‐rate samples (>2% missingness) and duplicated samples, the data set resulted in 83,717 unique imputed adult participants based on the eMERGE subject IDs from 77 imputation batches. The details of the imputation and quality control are given in [^9^](https://sciwheel.com/work/citation?ids=5917828&pre=&suf=&sa=0&dbf=0) .

**Methods**

**PRS-CSx**

We used PRS-CSx[^10^](https://sciwheel.com/work/citation?ids=12970124&pre=&suf=&sa=0&dbf=0), a recently developed Bayesian polygenic modeling method, to construct the ancestry-specific PRS while leveraging other ancestral GWAS summary statistics. PRS-CSx jointly models the GWAS summary statistics and couples genetic effects across populations using a shared continuous shrinkage prior, which enables more accurate effect size estimation by sharing information between summary statistics and leveraging linkage disequilibrium (LD) diversity across ancestral groups. The shared prior allows for correlated but varying effect size estimates across populations, retaining the flexibility of the modeling framework. We used pre-computed LD reference panels based on the UK Biobank data as provided with the PRS-CSx tool. The PRS-CSx included ~1.2 million HapMap3 variants in the PRS calculation.

**LDpred2**

LDpred2[^11^](https://sciwheel.com/work/citation?ids=10216417&pre=&suf=&sa=0&dbf=0) is an extension of the LDpred model which derives the PRS based on GWAS summary statistics and a matrix of correlation between genetic variants[^11^](https://sciwheel.com/work/citation?ids=10216417&pre=&suf=&sa=0&dbf=0). Here the summary statistics was obtained from the meta-analysis of the Asian and European GWAS. The correlation matrix was calculated as a weighted average of ancestry-specific LD matrices estimated based on a subset of Asian (nAsian = 7,370) and European (nEuropean = 61,493) samples, where the weight is proportional to the sample sizes of Asian and European GWAS in the summary statistics. We used the LDpred-grid model, which uses a grid of hyper parameters SNP heritability, ℎ^2^, and proportion of causal variants, *p*. LDpred2 estimates the heritability from constrained LD score regression. The heritability estimates for 1,020,293 SNPs retained in the PRS was 0.10. We used the validation data set, GERA cohort, to obtain the optimal value for the proportion of causal variants, *p* and heritability, ℎ^2^.

**PRS Calculation**

As described in [^12^](https://sciwheel.com/work/citation?ids=12898957&pre=&suf=&sa=0&dbf=0), the time that takes to calculate PRS may vary depending on the computation power, data set format and the tools selected for the analysis. We calculated the PRS using allelic dosages in PLINK 2[^13^](https://sciwheel.com/work/citation?ids=1158431&pre=&suf=&sa=0&dbf=0). Using nodes with ~300GB RAM for 20 cores, 200 SNPs took less than 24 milliseconds whereas 1 million SNPs took less than 72 milliseconds to generate PRS for each individual.

**Supplemental Table 1: Description of all studies included in the GWAS analysis -- European and Asian Summary Statistics (attached as separate excel sheet)**

**Supplemental Table 2: AUC estimates (95% confidence interval) for European-centric PRS, known loci PRS, PRS-CSx and LDpred2**

| **Case and Ethnicity** | **Studies** | **% of SNPs** | **n_case_/n_control_** | **EUR-centric PRS** | **Known Loci PRS** | **PRS-CSx** | **LDPred2** |
| --- | --- | --- | --- | --- | --- | --- | --- |
| **Asian** | GERA | 100% | 96/7,274 | 0.619  (0.547-0.689) | 0.587  (0.536-0.638) | 0.64  (0.58-0.69) | 0.627  (0.579-0.675) |
|  | MG-JPN | 100% | 2, 324/ 2,331 | 0.587  (0.571-0.603) | 0.601  (0.586-0.618) |  | 0.632  (0.616-0.644) |
| **Black or African American** | GERA | 100% | 56/3,103 | 0.526  (0.419-0.620) | 0.503  (0.444-0.613) |  | 0.546  (0.456-0.636) |
|  | MG-AA | 99% | 1856/ 4741 | 0.577  (0.560-0.596) | 0.580  (0.560-0.596) |  | 0.593  (0.576-0.612) |
|  | eMERGE^#^ | 100% | 42/4,025 | 0.574  (0.480-0.660) | 0.590  (0.500-0.690) |  | 0.620  (0.520-0.720) |
| **Latinx or Hispanic** | GERA | 100% | 70/6590 | 0.584  (0.511-0.656) | 0.587  (0.513-0.661) |  | 0.614  (0.547-0.681) |
|  | Hispanic GWAS | 100% | 1611/2106 | 0.612  (0.594-0.634) | 0.614  (0.592-0.632) |  | 0.635  (0.615-0.655) |
| **Non-Hispanic**  **White** | GERA | 100% | 1,401/75,611 | 0.638  (0.624-0.652) | 0.615  (0.600-0.629) | 0.64  (0.62-0.65) | 0.653  (0.638-0.668) |
|  | BCC | 100% | 873/945 | 0.617  (0.547-0.687) | 0.610  (0.550-0.670) |  | 0.645  (0.580-0.710) |
|  | CPS II | 100% | 804/908 | 0.619  (0.591-0.647) | 0.603  (0.576-0.630) |  | 0.634  (0.606-0.6624) |
|  | eMERGE^#^ | 99.7% | 573/37,641 | 0.620  (0.600-0.640) | 0.610  (0.54-0.68) |  | 0.640  (0.610-0.670) |
|  | *^ Number of Variants Available / Number of Variants in Score (%) of LDpred2*  *# AUC is adjusted for age, sex and 10 PCs* | | | | | | |

**Supplemental Table 3**: Odds Ratios (OR) and 95% confidence interval (95% CI) for PRS per SD and stratified by family history and age.

|  | **Asian** | | **Black or African American** | | **Latinx or /Hispanic** | | **Non-Hispanic White** | | |
| --- | --- | --- | --- | --- | --- | --- | --- | --- | --- |
|  | ***GERA*** | **MG-JPN** | ***GERA*** | **MG-AA** | ***GERA*** | ***Hispanic GWAS*** | ***GERA*** | ***BCC*** | ***CPS II*** |
|  | **OR (95% CI)**  **p-value** | **OR (95% CI)**  **p-value** | **OR (95% CI)**  **p-value** | **OR (95% CI)**  **p-value** | **OR (95% CI)**  **p-value** | **OR (95% CI)**  **p-value** | **OR (95% CI)**  **p-value** | **OR (95% CI)**  **p-value** | **OR (95% CI)**  **p-value** |
| **PRS per SD** | 1.58  (1.29-1.94)  1.3e-05 | 1.64  (1.55-1.73)  <2.0e-18 | 1.25  (0.96-1.64)  0.090 | 1.40  (1.33-1.48)  <2.0e-18 | 1.45  (1.16-1.80)  1.3e-03 | 1.63  (1.52-1.75)  <2.0e-18 | 1.70  (1.62-1.79)  <2.0e-18 | 1.72  (1.43-2.07)  7.93e-09 | 1.58  (1.44-1.74)  <2.0e-18 |
| **Family History** | | | | | | | | | |
| **No** | 1.58  (1.24-2.00)  1.8e-04 | 1.64  (1.46-1.83)  7.00e-17 | 1.30  (0.97-1.1.73)  0.08 | 1.73  (1.34-2.22)  2.03e-05 | 1.53  (1.20-1.96)  6.7e-04 | 1.65  (1.52-1.79)  <2.0e-18 | 1.69  (1.60-1.79)  <2.0e-18 |  | 1.60  (1.44-1.78)  2.00e-18 |
| **Yes** | 1.30  (0.84-2.01)  0.229 | 1.49  (1.20-1.84)  3.4e-04 | 1.05  (0.535-2.05)  0.89 | 1.23  (1.04-1.46)  1.23e-02 | 0.998  (0.54-1.82)  0.995 | 1.18  (0.91-1.53)  0.206 | 1.54  (1.38-1.72)  4.58e-14 |  | 1.34  (1.00-1.77)  4.55e-02 |
| **Age** | | | | | | | | | |
| ***<50*** | 13 cases and 1,295 controls | 174 cases and 166 controls | 4 cases and 382 controls | 293 cases and 98 controls | 13 cases and 1288 controls | 258 cases and 2 controls | 59 cases and 5,127 controls | 20 cases and 676 controls | -- |
|  | 1.69  (0.954- 2.98)  0.07 | 1.92  (1.50- 2.45)  2.13e-07 | 1.36  (0.484- 3.84)  0.56 | 1.52  (1.17-1.97)  1.58e-03 | 1.17  (0.683- 2.00)  0.569 | -- | 1.93  (1.50- 2.48)  3.22e-07 | 1.60  (1.00- 2.54)  0.04 |  |
| ***50 to 60*** | 29 cases and 1,390 controls | 516 cases and 494 controls | 12 cases and  539 controls | 478 cases 822 controls | 14 cases and 1318 controls | 434 cases and198 controls | 196 cases and 9,743 controls | 91 cases and 212 controls | 41 cases and 50 controls |
|  | 1.68  (1.14-2.48)  0.009 | 1.87  (1.62-2.16)  1.17e-17 | 1.51  (0.824-2.76)  0.182 | 1.53  (1.36-1.72)  2.11e-12 | 1.80  (1.08-2.99)  0.023 | 2.22  (1.80-2.74)  3.52e-16 | 1.80  (1.57- 2.06)  1.41e-17 | 1.58  (1.22-2.06)  6.01e-04 | 1.72  (1.02-2.88)  0.042 |
| ***60 to 70*** | 21 cases and 2,064 controls | 832 cases and 877 controls | 19 cases and 872 controls | 630 cases and 1,661 controls | 17 cases and 1,710 controls | 495 cases and 990 controls | 377 cases and 19,831 controls | 213 cases and 56 controls | 301 cases and 337 controls |
|  | 1.63  (1.06- 2.50)  0.024 | 1.57  (1.42- 1.74)  <2.0e-18 | 1.87  (1.145-3.06)  0.012 | 1.40  (1.27-1.54)  1.88e-11 | 1.67  (1.03- 2.69)  0.036 | 1.58  (1.42-1.77)  3.81e-16 | 1.92  (1.74- 2.12)  <2.0e-18 | 2.18  (1.49- 3.20)  6.69e-05 | 1.72  (1.46-2.01)  3.05e-11 |
| ***70 to 80*** | 21 cases and 1,437 controls | 652 cases and 693 controls | 15 cases and  806 controls | 380 cases and 1,824 controls | 17 cases and 1,379 controls | 329 cases and 760 controls | 426 cases and  21,932 controls | -- | 346 cases and 398 controls |
|  | 1.52  (0.955- 2.40)  0.077 | 1.57  (1.40- 1.76)  6.91e-15 | 0.84  (0.51-1.38)  0.494 | 1.33  (1.19- 1.549)  3.46e-07 | 1.252  (0.768- 2.04)  0.367 | 1.49  (1.30-1.69)  1.78e-09 | 1.66  (1.52-1.82)  <2.0e-18 | -- | 1.49  (1.29-1.72)  8.24e-08 |
| ***>80*** | 12 cases and 1,088 controls | 101 cases and 150 controls | 6 cases and  504 controls | 75 cases and 336 controls | 9 cases and 895 controls | 95 cases and 156 controls | 343 cases and 18,978 controls | -- | 116 cases and 123 controls |
|  | 1.27  (0.682- 2.38)  0.477 | 1.74  (1.31- 2.30)  1.08e-04 | 0.87  (0.36-2.06)  0.745 | 1.37  (1.05- 177)  1.86e-02 | 1.5864  (0.83- 2.99)  0.16 | 1.57  (1.18-2.10  2.0e-03 | 1.42  (1.28- 1.58)  1.90e-11 |  | 1.50  (1.15-1.95)  2.79e-03 |

**Supplemental Table 4: SNP associations for newly and previously identified colorectal cancer risk loci (attached as separate excel sheet)**

**Supplemental Figure 1:** ROC curves of PRS for CRC, stratified by race and ethnicity

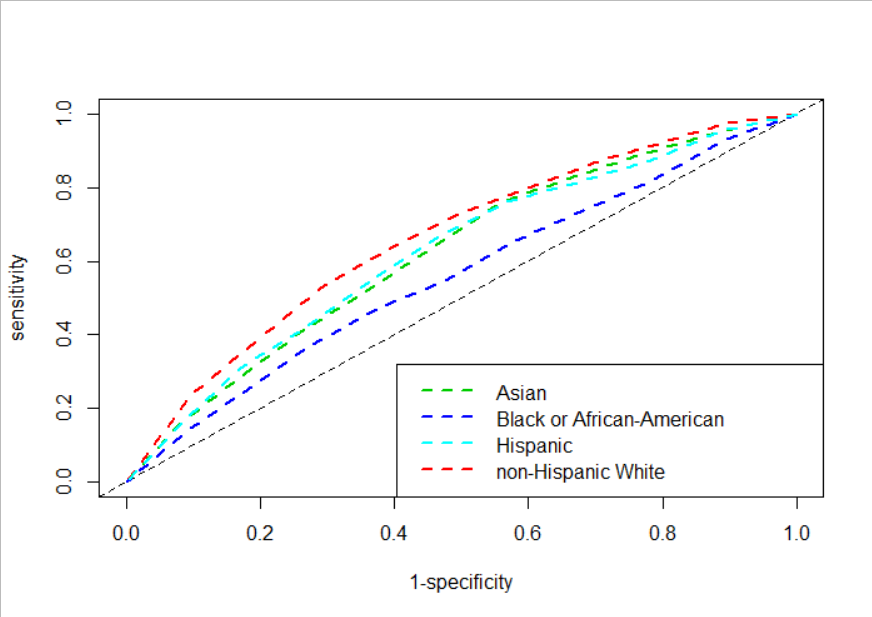

**Supplemental Figure 2:** Distribution of PRS for GERA cohort. A) PRS distributions varied across the racial and ethnic groups, B) PRS distribution after ancestry adjustment.

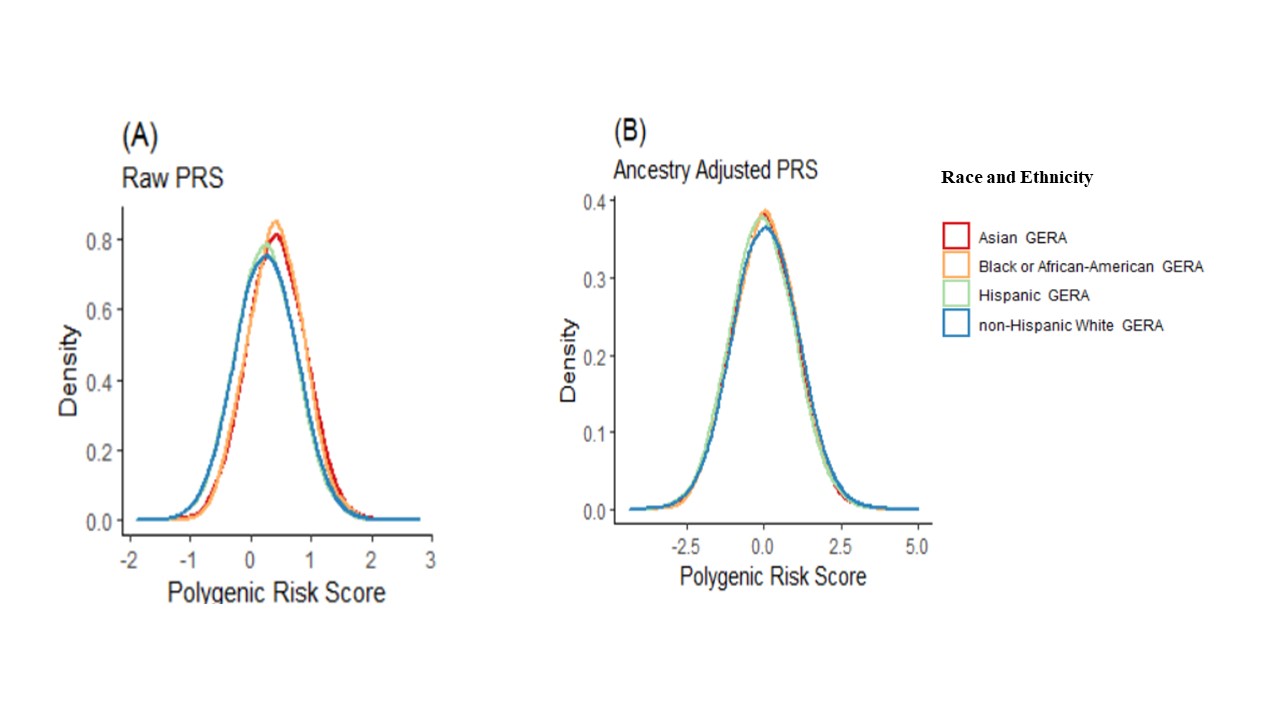

**Supplemental Figure 3:** Distribution of PRS for all other validation studies except for the GERA cohort. A) PRS distributions varied across the racial and ethnic groups, B) PRS distribution after ancestry adjustment, C) Additional mean adjustment for the MG-JP that has a different imputation panel.

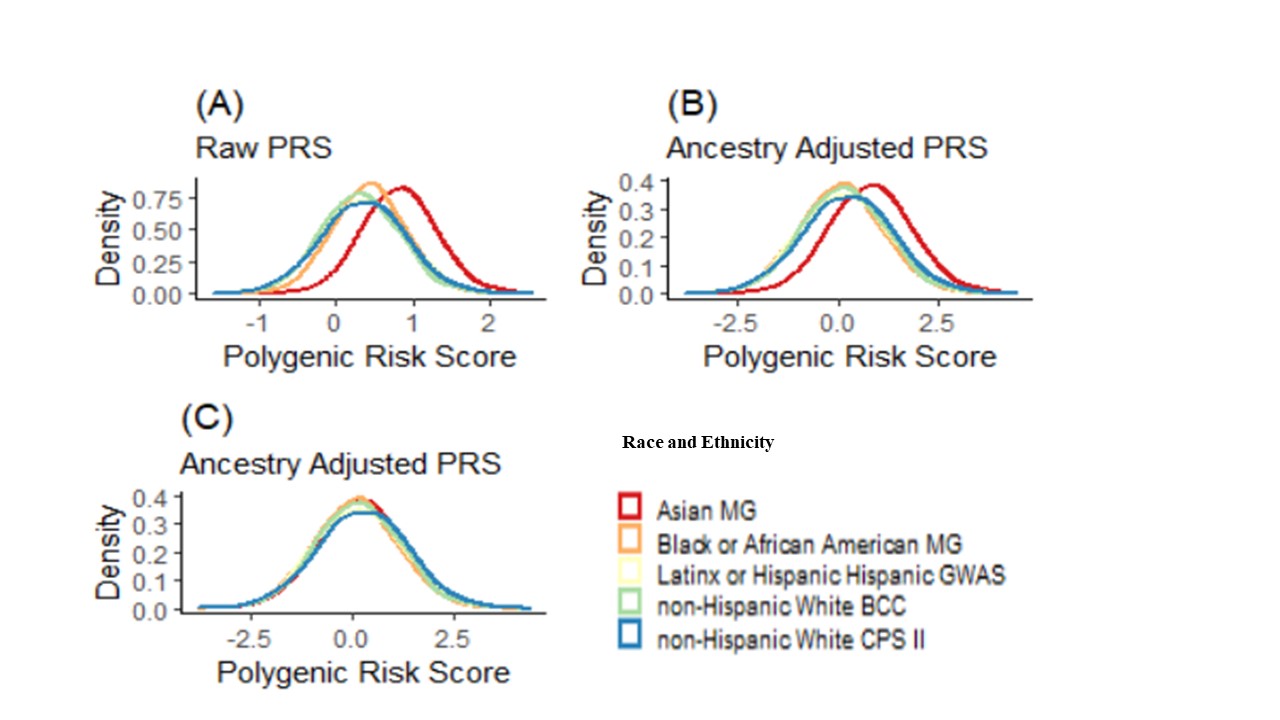

**Supplemental Figure 4:** Distribution of the ancestry adjusted PRS by race and ethnicity and by colorectal cancer cases and controls. PRS is the single cross-ancestry Asian-European PRS based on LDpred2.

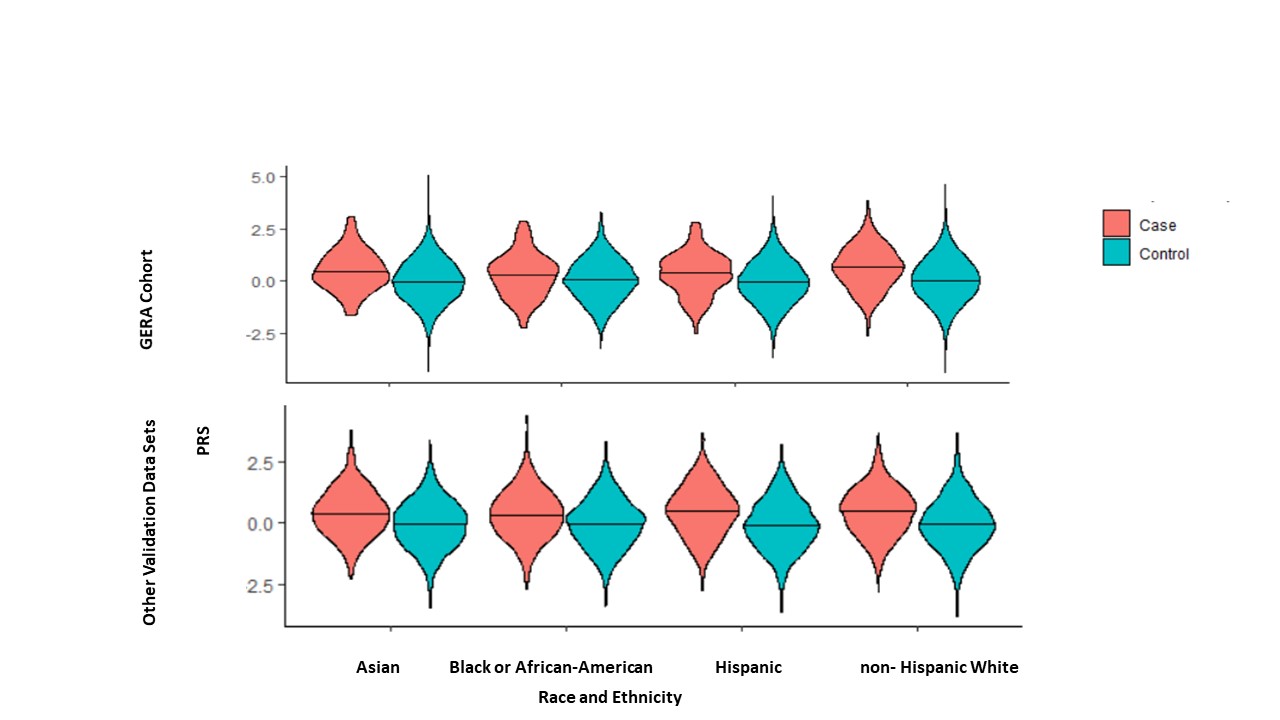

**Supplemental Figure 5:** Decision-Curves for the model based on family history and PRS, model based on family history alone, intervene for all, and intervene for none, in GERA study participants with age 50-59, stratified by ancestral groups.

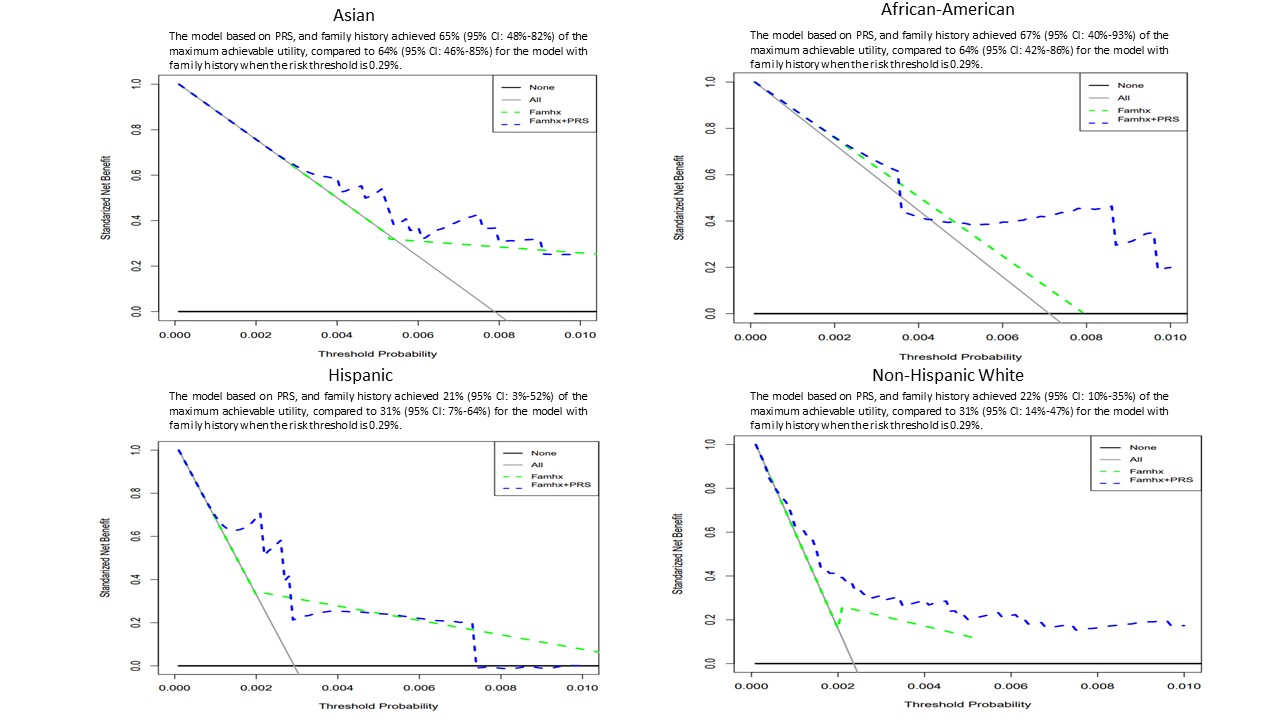

**Supplemental Figure 6:** Decision-Curves for the model based on family history and PRS, model based on family history alone, intervene for all, and intervene for none, in GERA study participants with age 50-59. The model based on PRS, and family history achieved 52.4% (95% CI: 45%-60%) of the maximum achievable utility, compared to 45.1% (95% CI: 35%-54%) for the model with family history only (p-value = 0.045) when the risk threshold is 0.29%.

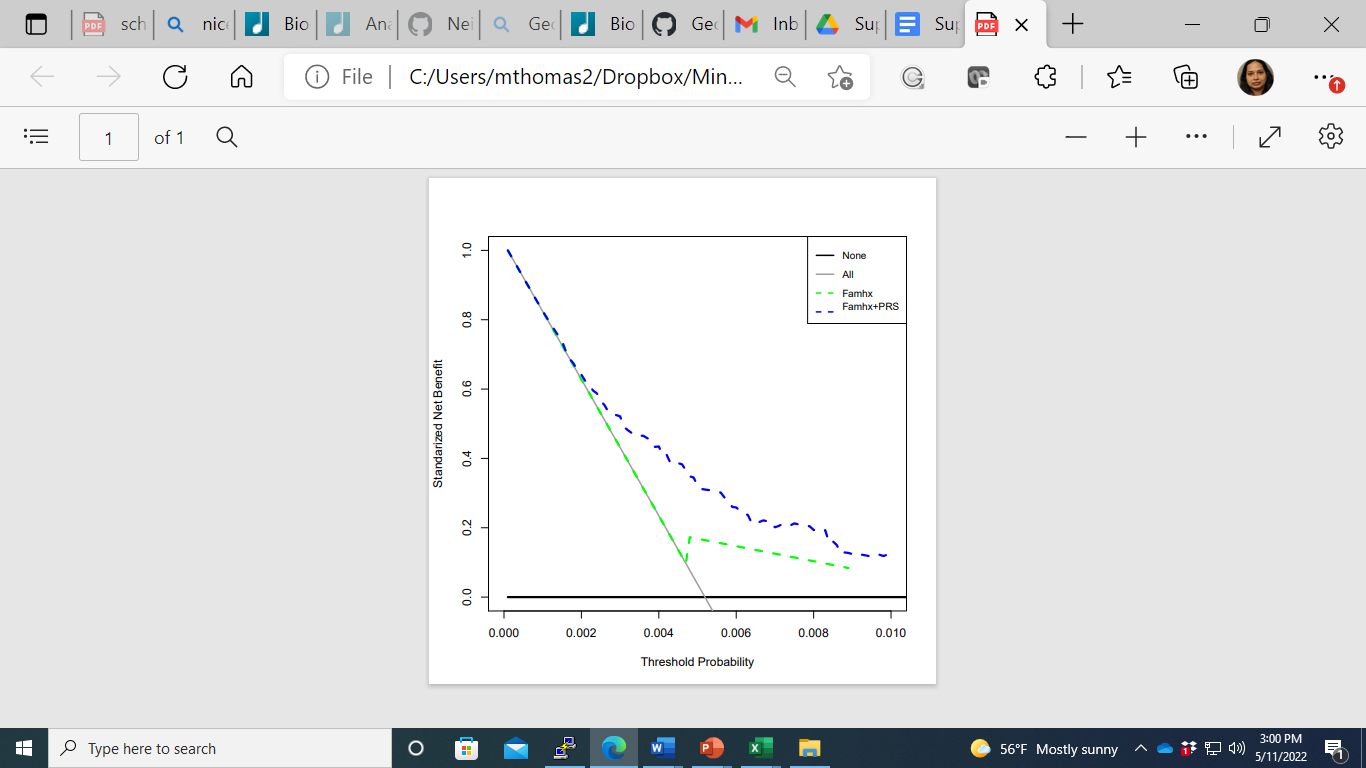
